# MDSGene update and expansion: Clinical and genetic spectrum of *LRRK2* variants in Parkinson’s disease

**DOI:** 10.1101/2024.12.10.24318787

**Authors:** Clara Krüger, Shen-Yang Lim, Alissa Buhrmann, Fenja L. Fahrig, Carolin Gabbert, Natascha Bahr, Harutyun Madoev, Connie Marras, Christine Klein, Katja Lohmann

## Abstract

Pathogenic variants in the *LRRK2* gene are one of the most commonly identifiable monogenic causes of Parkinsońs disease (PD, PARK-LRRK2). This systematic MDSGene literature review comprehensively summarizes published demographic, clinical, and genetic findings related to potentially pathogenic *LRRK2* variants (https://www.mdsgene.org/). Recent insights on LRRK2’s kinase activity have been incorporated for pathogenicity scoring.

Data on 7,885 individuals with 292 different variants were curated, including 3,296 patients with PD carrying 205 different potentially disease-causing *LRRK2* variants. The initial MDSGene review covered only 724 patients carrying 23 different *LRRK2* variants. Missingness of phenotypic data in the literature was high, hampering the identification of detailed genotype-phenotype correlations. Notably, the median age at onset in the patients with available information was 56 years, with approximately one-third having PD onset <50 years. Tremor was the most frequently reported initial symptom and more frequent than reported in other dominantly inherited forms of PD. Of the 205 potentially disease-causing variants, 14 (6.8%) were classified as pathogenic, 8 (3.9%) as likely pathogenic, and the remaining 183 (89.3%) as variants of uncertain significance (VUS). The pathogenic p.G2019S variant was the most frequent pathogenic variant, followed by p.R1441G and p.R1441C, accounting for >80% of patients, with Tunisia, Spain, and Italy contributing about half of patients.

This systematic review represents the largest database on PARK-LRRK2 to date and provides an important resource to improve precision medicine. Given their high frequency, a better interpretation of the pathogenicity of VUS is needed for selection and stratification of patients in clinical trials.

## Introduction

Parkinson’s disease (PD) is a relatively common, age-related, neurodegenerative disorder characterized by motor (bradykinesia, resting tremor, rigidity, and postural instability) and non-motor features (neuropsychiatric features, autonomic symptoms, sleep disorders, and sensory dysfunction).^1^ The age at onset for almost 25% of affected individuals is younger than 65 years, and for 5–10%, younger than 50 years.^2^ Causes of PD include genetic and environmental factors as well as interactions thereof.^2^ Monogenic causes, i.e., pathogenic variants in genes such as *LRRK2, SNCA, VPS35, PRKN, PINK1,* and *PARK7,* often play a role in patients with early-onset disease.^3, 4^ Further, risk variants and rare pathogenic variants (often with highly reduced penetrance)^5^ in the *GBA1* gene are found in about 10% of PD patients.^6, 7^ Although monogenic forms comprise a minority of all PD patients^6–8^, they are important as affected individuals are the most likely candidates for potential gene-specific, targeted treatments. These treatments are currently being evaluated in trials^2^, including kinase inhibitors for PD linked to pathogenic variants in the *LRRK2* gene (PARK-LRRK2).^9^

Pathogenic variants in the *LRRK2* gene are one of the most frequent causes of dominantly inherited PD.^10^ *LRRK2* (Leucine-Rich Repeat Kinase 2) encodes a multidomain protein kinase that shows autophosphorylation.^11^ It plays an important role in different cellular processes, such as cytoskeleton remodeling, vesicular trafficking, autophagy, and protein translation.^12^ Increased LRRK2 kinase activity is thought to dysregulate these processes resulting in the death of dopaminergic neurons in the *substantia nigra*.^13^ Until recently, only seven variants in *LRRK2* were considered clearly pathogenic.^4, 14, 15^ These variants are mainly located in the guanine triphosphatase (GTPase) Ras-of-complex (ROC) and the kinase domain, which represent two functionally linked enzymatic domains.^10, 16^ More recently, in-vivo and in-vitro functional testing of LRRK2’s kinase activity became available and revealed approximately 20 additional variants that lead to increased LRRK2 kinase activity and are thus plausible contributors to the pathogenesis of PD.^16, 17^ Importantly, the increased LRRK2 kinase activity represents a possible therapeutic target.^9, 18^ In addition to these pathogenic kinase-activating, gain-of-function variants in *LRRK2*, there are also loss-of-function variants, i.e., nonsense and frameshift variants. However, these truncating variants in *LRRK2* are found with similar frequency in PD patients and controls, indicating that *LRRK2* loss-of-function variants may not alter the risk for PD.^19, 20^ Further, copy number variants (CNVs), especially larger deletions, are extremely rarely found in *LRRK2*^21, 22^.

The very large number of scientific publications related to PARK-LRRK2 (>3,000), makes it overwhelming and challenging to follow the literature. Despite the large body of literature, there remain many uncertainties, especially with respect to the phenotypic spectrum and the interpretation of the many genetic variants. These knowledge gaps potentially hamper the identification of PARK-LRRK2 patients who would be eligible for gene-specific clinical trials. Here, we provide a systematic literature review using the protocol of the Movement Disorders Society Genetic Mutation Database (MDSGene) (https://www.mdsgene.org/).^23^ With this review, we update and considerably extend the MDSGene database to over 200 potentially disease-causing *LRRK2* variants reported in more than 3,000 PD patients. The numbers of included variants and patients have increased approximately tenfold and fourfold, respectively, compared to the initial review covering publications until 2017.^4^

## Materials and Methods

### Literature search and Data Collection Process

For the literature search, we used the PubMed database (https://pubmed.ncbi.nlm.nih.gov/). The search term followed the MDSGenés format (Suppl. Table 1), and articles were screened stepwise based on title, abstract, and full text.^24^ We included articles that reported at least one individual with a *LRRK2* variant that was considered potentially disease-related by the authors. Further, we evaluated the literature cited in the included articles and used the Human Gene Mutation Database (HGMD) professional (https://apps.ingenuity.com/ingsso/login)^25^ to identify additional eligible papers.

Demographic, genetic, and clinical information from all eligible papers were extracted using the MDSGene protocol.^4^ Evaluated variables are listed in Suppl. Table 2.

### Inclusion and Exclusion Criteria for Patients and Genetic Variants

We only included patients with PD and a potentially pathogenic *LRRK2* variant. Unaffected mutation carriers, patients with atypical parkinsonian disorders (dementia with Lewy bodies [DLB], multiple system atrophy [MSA], progressive supranuclear palsy [PSP], and corticobasal degeneration/syndrome [CBD/CBS]), and patients with non-movement disorder conditions were also extracted but excluded from being displayed on the MDSGene database and from the analyses for PD-related genotype-phenotype correlations. Where authors and patient details suggested duplicate reporting of the same individual, we combined information to create a single entry. Patients who carried additional potentially PD-causing variants were also excluded. Although *LRRK2* is inherited in an autosomal dominant manner, patients with heterozygous and homozygous variants were included as before^4^ since previous literature indicated that there is no dosage effect.^26^ We excluded *LRRK2* variants that had a minor allele frequency (MAF) of >1% in any ethnicity reported in the gnomAD Browser (https://gnomad.broadinstitute.org/). Carriers of benign or likely benign *LRRK2* variants were also excluded.

### Pathogenicity Scoring

All variants were mapped to GRCh37/hg19, and the nomenclature is based on the transcript ENST00000298910 for *LRRK2*. For pathogenicity scoring, we followed the recommendations of the American College of Medical Genetics and Genomics (ACMG).^27^ For this, we used two publicly available online tools, i.e., VarSome (https://varsome.com/) and Franklin (https://franklin.genoox.com/). If there was a discrepancy between the two tools or the score was not plausible, variants were manually scored according to the ACMG recommendations.^27^ For this, we applied the CADD score^28^ as an in-silico measurement and data from the functional testing for 100 *LRRK2* variants.^16^ Variants categorized as “pathogenic”, “likely pathogenic”, or “variant of uncertain significance” (VUS) were considered as potentially disease-causing variants and included in the MDSGene database and the analyses.

## Results

### Included Articles and Study Types

We identified 3,064 publications through the search strategy, carried out for the last time in PubMed on January 10^th^, 2024, and supplemented with articles from HGMD professional v2022.4. An overview of the literature search and the filtering process is shown in Figure 1. At screening, 880 articles were considered eligible for data abstraction. However, 521 did not contain relevant information (Figure 1). The full text of 359 papers was reviewed, but only 291 publications included patients who fulfilled the inclusion criteria (Figure 1).

**Figure 1:**
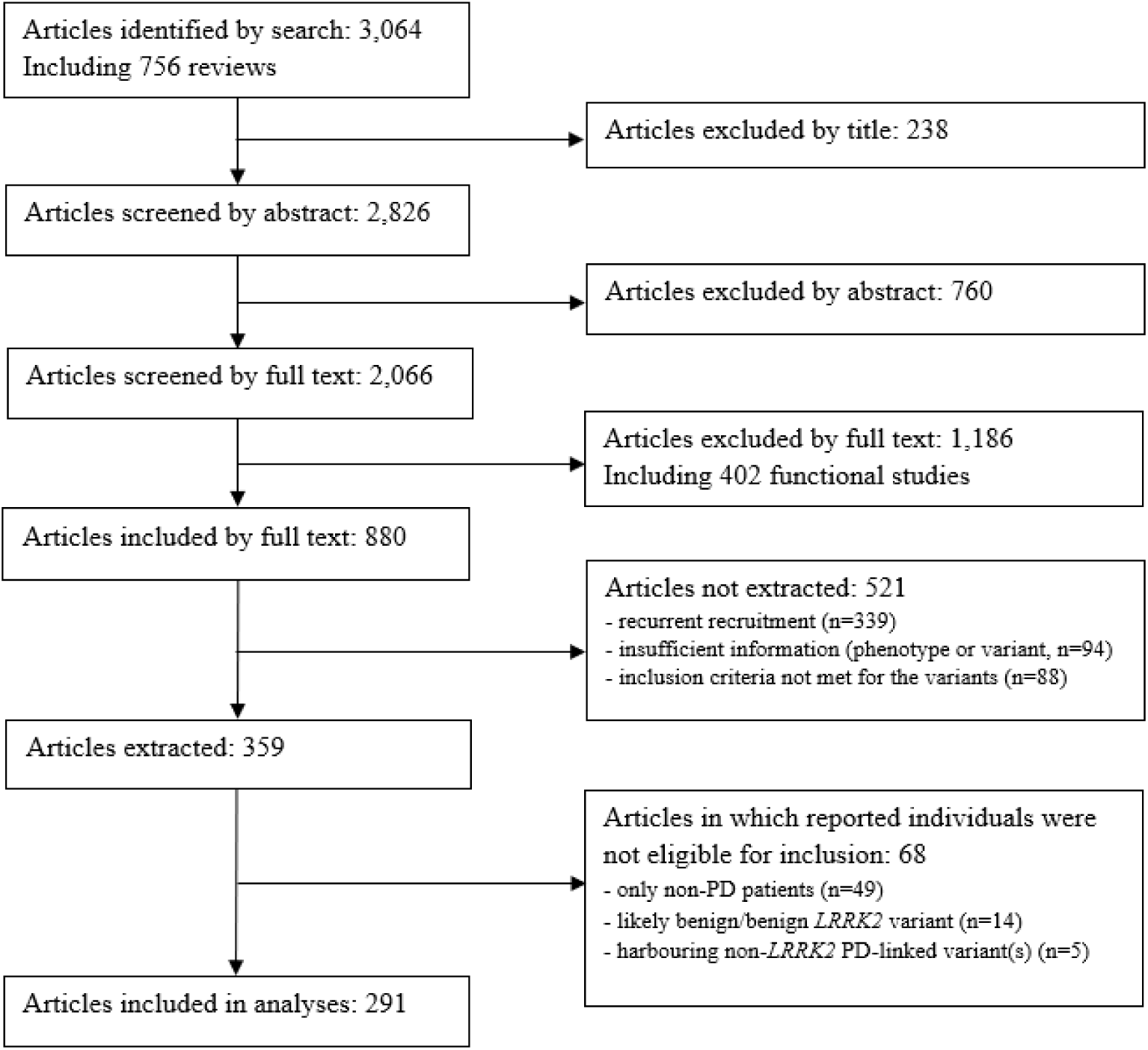
Flowchart of the literature search. The number of included and excluded articles at the different steps is indicated.

The most frequently included study types were mutational screens (161/291, 55.3%), followed by case reports or case series (46/291, 15.8%), association studies (35/291, 12.0%), family studies (16/291, 5.5%), and a combination of different designs (33/291, 11.3%).

### Pathogenicity Scoring

A total of 292 *LRRK2* variants were identified on the DNA level corresponding to 291 variants on the protein level since there were two changes (c.5385G>C and c.5385G>T) resulting in the same amino acid substitution (p.L1795F). Of the 292 variants, 290 were scorable using VarSome and Franklin. The remaining two variants and 176 of the 290 variants were manually scored following the ACMG recommendations^27^. There were considerable discrepancies in the pathogenicity scoring between VarSome and Franklin (Figure 2). The manual scoring was closer to Franklin (Figure 2). One reason for the discrepancies was an underestimation in VarSome, which interpreted many VUS erroneously as “likely benign” since the criterion BP1 was applied (“Missense variant in a gene for which primarily truncating variants are known to cause disease”). This is not applicable to *LRRK2*, since loss of function of the protein is not a known disease mechanism.^19, 20^ In fact, many missense variants in *LRRK2* have instead been demonstrated to be functionally relevant variants^16^. Using BP1 resulted in scoring almost half of the variants as likely benign by VarSome. Another discrepancy can be explained by an underestimation of likely pathogenic variants, being classified as VUS, in Franklin since the criterion PS3 was not applied (“Well-established in-vitro or in-vivo functional studies supportive of a damaging effect on the gene or gene product.”), even though functional evidence for these variants has become available^16^. In the combined and finally considered classification, the vast majority of the variants were classified as VUS (240/292, 82.2%; Figure 2). A list of all *LRRK2* variants can be found in Suppl. Table 3.

**Figure 2:**
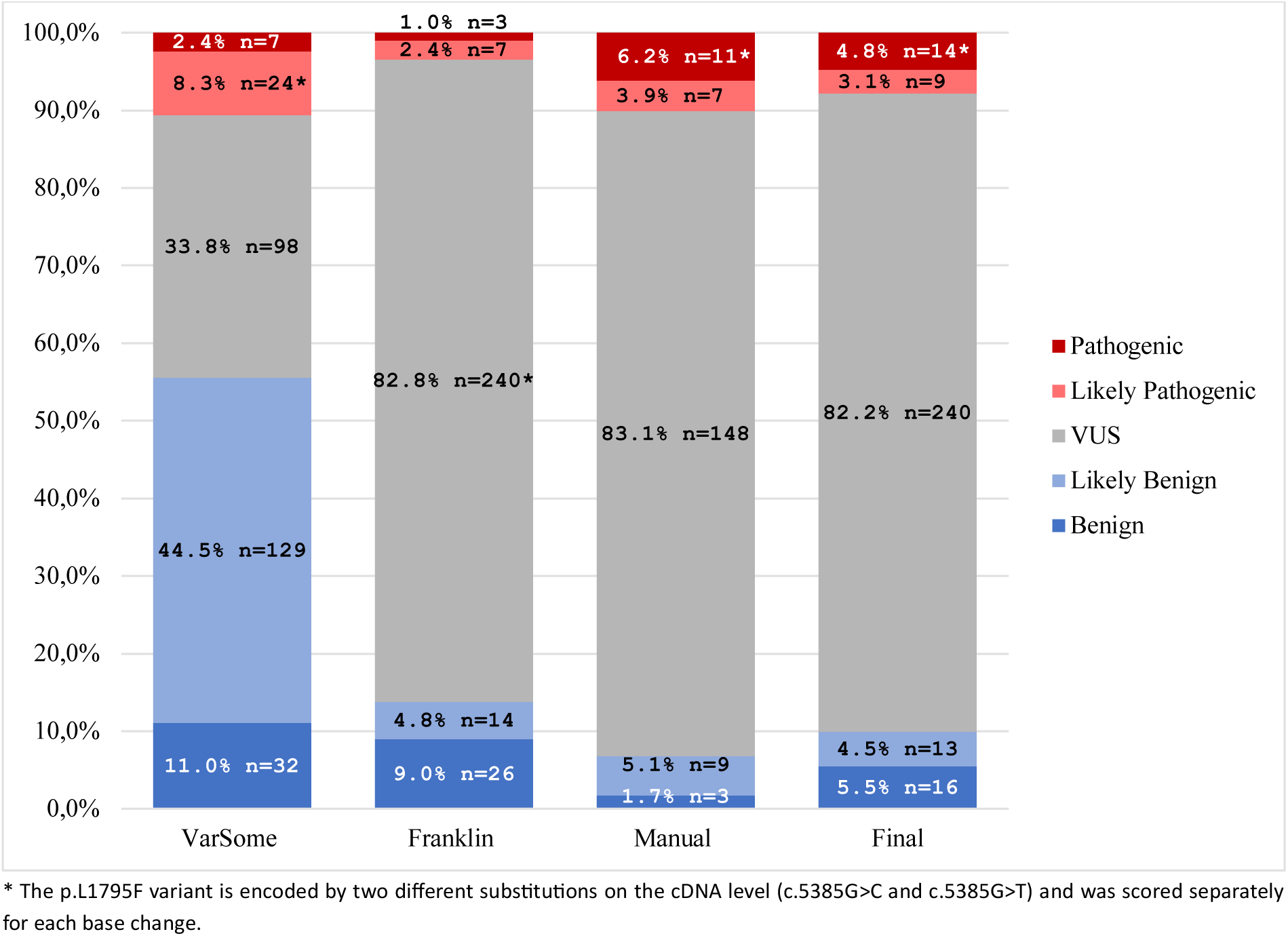
Results of the pathogenicity scoring. Comparison of the ACMG-based pathogenicity scoring of all *LRRK2* variants from VarSome, Franklin, manual ACMG pathogenicity scoring, and the final pathogenicity scoring. * The scoring was done on the cDNA level. Therefore, the two variants encoding p.L1795F were counted separately.

### Included Patients and Variants

A total of 7,885 *LRRK2* variant carriers were identified, and 3,296 PD patients with a potentially pathogenic *LRRK2* variant (41.8%) were included in MDSGene and the analyses, based on the inclusion and exclusion criteria. Among the 7,885 individuals, 75.7% (n=5,969) of them had PD, 1.8% (n=138) had other diseases (see below), and 22.5% (n=1,778) were clinically unaffected. The included 3,296 PD patients carried 205 different variants, of which 14 have been classified as pathogenic (6.8%), 8 as likely pathogenic (3.9%), and 183 as VUS (89.3%; Figure 3).

**Figure 3:**
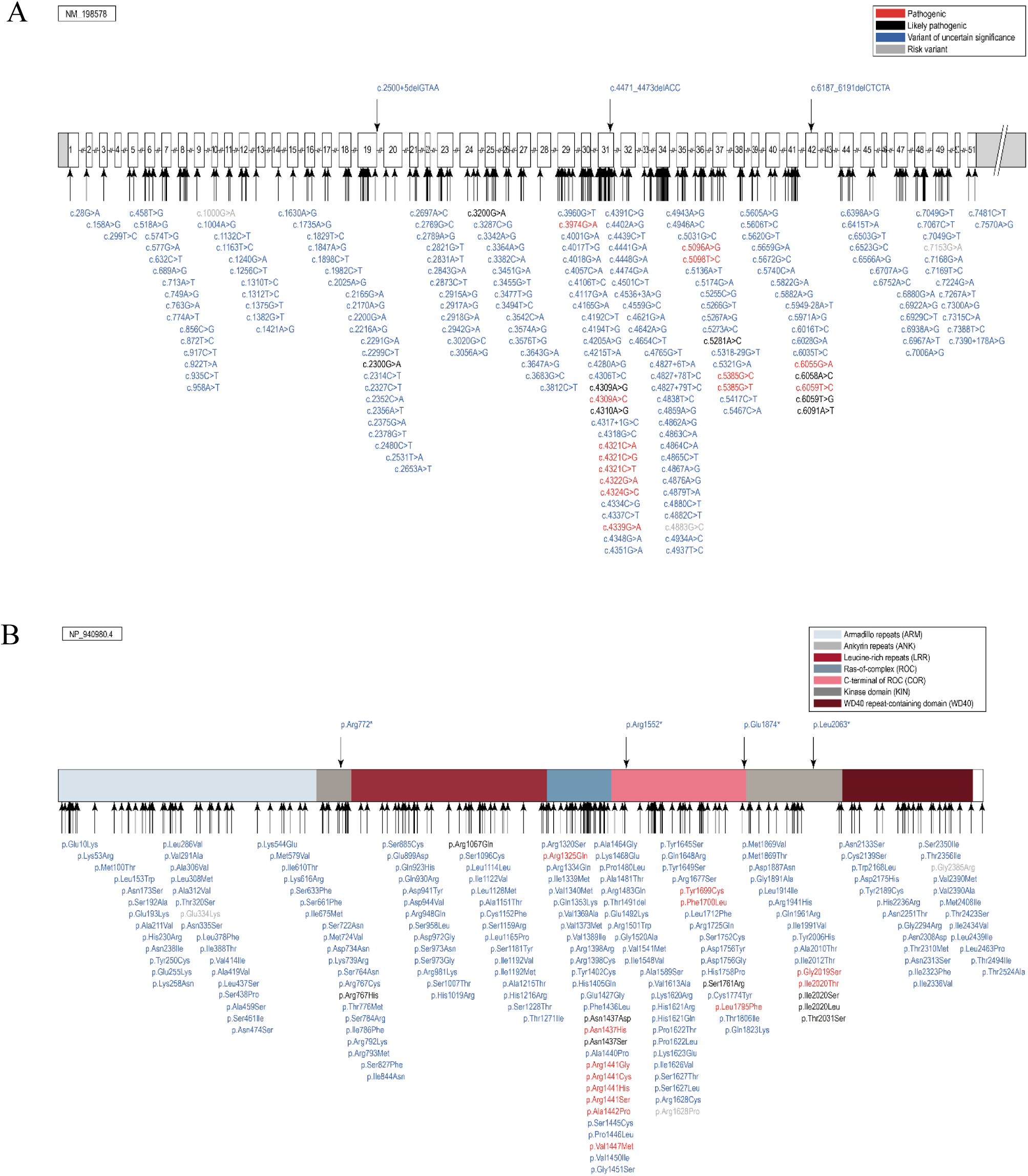
Potentially pathogenic variants in *LRRK2*. Schematic representation of the *LRRK2* gene **(A)** and protein **(B)** with the localization of the included variants. The position of the variants is indicated by arrows, and the predicted pathogenicity is provided by color (red, pathogenic; black, likely pathogenic; blue, VUS; grey, risk variant).

Of the 205 included variants, missense variants (91.7%, n=188) were the most frequent variant type, followed by intronic variants (2.4%, n=5), nonsense variants (2.0%, n=4), splice region variants (2.0%, n=4), silent changes (1.0%, n=2), splice site variants (0.5%, n=1), and small deletions (0.5%, n=1). Of note, all variants other than missense variants were classified as VUS. The most frequent variant among the included patients was the p.G2019S substitution, classified as pathogenic, that was present in 2,426 patients (73.6%). Notably, among the clinically unaffected individuals, 739 (41.6%) carried this missense variant, i.e., 739 of 3,165 (23.3%) p.G2019S carriers were unaffected (age at examination unknown for 640 of the unaffected patients, 86.6%). The second most frequently reported variant among the included patients was p.R1441G in 193 patients (5.9%). This variant was also reported in 90 unaffected individuals, which means that 90 of the 283 (31.8%) reported carriers were unaffected (Suppl. Table 4). Since unaffected variant carriers are not systematically reported and have not been extracted from the beginning of the project^4^, no definite conclusion on the penetrance of these variants can be drawn. Nevertheless, incomplete penetrance for p.G2019S and also for p.R1441G plays a considerable role since a large proportion of individuals with these variants had not developed manifest PD at the point of inclusion.

### Excluded Patients and Variants

Notably, among the 7,885 individuals with *LRRK2* variants, 0.3% (n=25) had an atypical parkinsonian disorder, and 1.4% (n=113) had other diseases (such as immunological diseases, cancer, dementia, or essential tremor). These were excluded from the analyses since the sole causative role of the *LRRK2* genotype in these non-PD phenotypes has not been established.

Among the 138 patients with a non-PD phenotype extracted during the systematic literature review, 9 patients also carried a potentially disease-causing variant in another gene. Of the remaining 129 patients, only 20 carried pathogenic or likely pathogenic variants, mostly p.G2019S, 59 carried VUS, and 50 benign or likely benign variants. The phenotypic spectrum of carriers with (likely) pathogenic variants was broad, including MSA (n=3)^29^ ^30^, PSP (n=3)^31–33^, CBD (n=1)^34^, dystonia (n=2)^35^ ^36^, Alzheimeŕs disease (n=1)^37^, amyotrophic lateral sclerosis (n=2)^38^, restless legs syndrome (n=1)^39^, schizophrenia (n=1)^40^, multiple sclerosis (n=3)^41^, rheumatoid arthritis (n=1)^41^, achalasia (n=1)^41^, and breast cancer (n=1)^42^. Often, these non-PD phenotypes were found in relatives of PD patients with the same *LRRK2* variant. Of note, some of these phenotypes are frequent. Thus, it is also conceivable that there was a co-occur by chance, and these patients show reduced penetrance of the *LRRK2* variant in terms of PD.

Regarding the 25 patients with atypical parkinsonian disorders, 7 patients carried pathogenic *LRRK2* variants (see above), 11 carried VUS, and 7 had benign or likely benign *LRRK2* variants. Among the 18 carriers with potentially disease-causing *LRRK2* variants were 8 MSA, 7 PSP, 2 CBD, and 1 DLB patient(s).

In addition to the rare, presumably monogenically acting variants, three additional coding variants should be mentioned that may increase the risk of developing PD. These include two variants (p.G2385R and p.R1628P) that have repeatedly been shown to act as risk factors for PD in Asian populations.^43, 44, 45^ The third variant, p.E334K, is frequently observed in the Finnish population and showed increased LRRK2 kinase activity.^16^ These variants have not been included in MDSGene, since their MAF is >1% in certain populations.

We further excluded 44 patients whose genetic cause could not be unequivocally assigned to *LRRK2* since they carried potentially pathogenic variants in at least one other PD gene. This included 18 patients with an additional, potentially disease-causing variant in *GBA1*, and 13 patients with additional *PRKN* variants. Notably, one patient, who, to our knowledge, is the only case in the literature carrying a large deletion in *LRRK2* (homozygous Exon 49deletion), also carried a homozygous, pathogenic deletion of Exon 4 in *PRKN*.^22^

### Demographic and Phenotypic Data

One of the biggest challenges for this systematic review was the missing phenotypic data in the majority of publications. Missing data in up to 99.9% of the patients were observed for some of the extracted variables (Figure 4). Even for the four cardinal clinical features of PD, the range of missing data was from 25.3% for bradykinesia to 89.0% for postural instability (Figure 4A).

**Figure 4:**
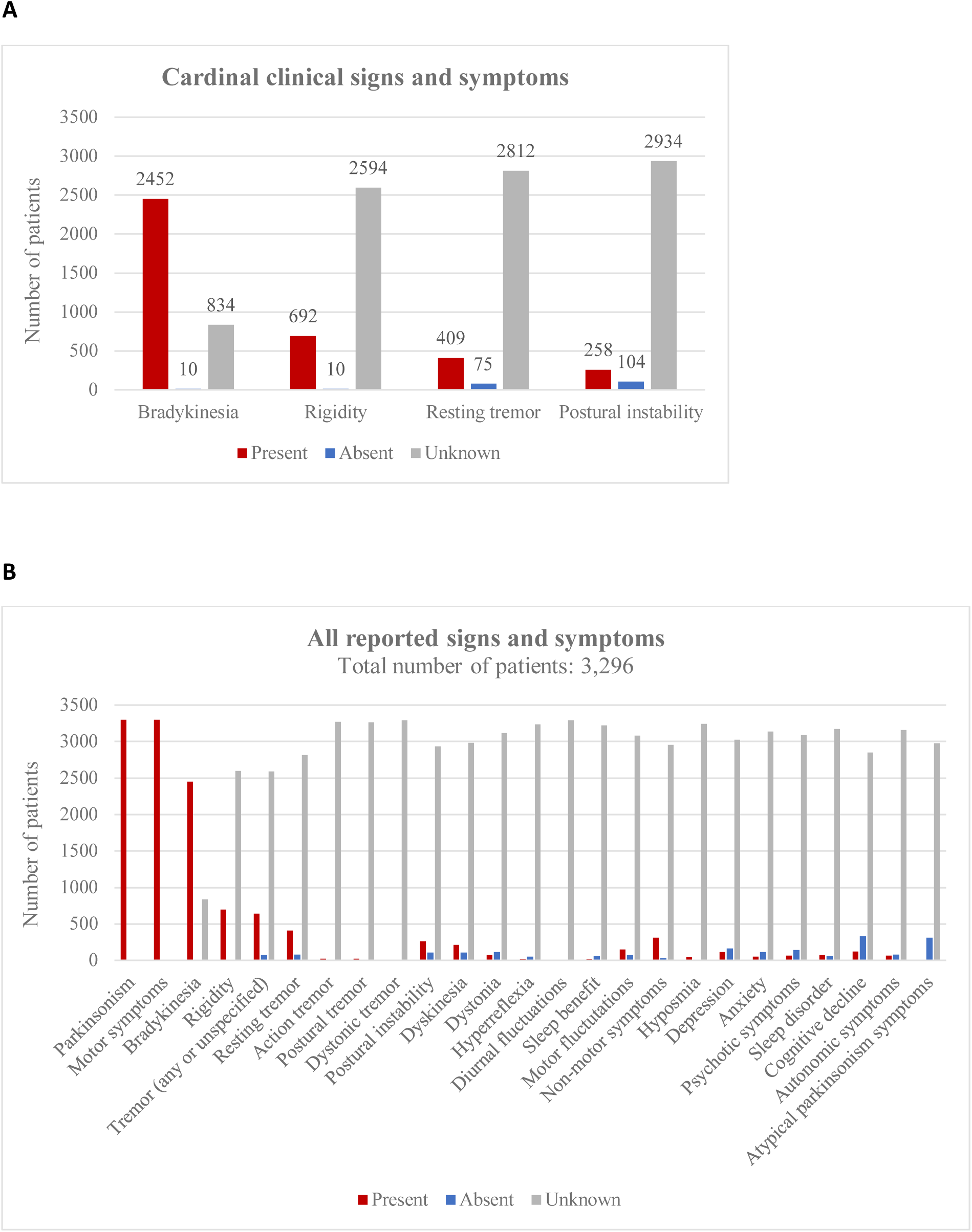
Signs and symptoms in the included patients at last examination. **A)** Cardinal clinical signs and symptoms in the included PARK-LRRK2 patients. **B)** Overview of all reported signs and symptoms in the included patients.

Among the 3,296 included patients, information on age was available for 762 patients (23.1%) who had a median age of 67 (range 26-95, interquartile range: 58-75) years (the mean age with standard deviation was 65.9±12.2 years). Information on sex was not provided for 1,847 patients (56.0%), among the remaining 1,449 patients, 758 (52.3%) were male. Patients were most commonly European/White (32.8%), Arab (24.1%), or of mixed/other ethnicities (14.9%), based on 1,647 patients with available information (50.0% missing data; Suppl. Figure 1A). Most reported patients originated from Tunisia (26.3%), Spain (14.9%), Italy (6.4%), China (5.9%), or the USA (5.3%) (Suppl. Figure 1B). Notably, among the Tunisian patients, almost all (98.6%, 545/553) carried the p.G2019S variant (median AAO 58 years), while in Spain, about half of the patients carried this variant (47.3%, 148/313, median AAO 62 years) and the other half the p.R1441G variant (49.5%, 155/313, median AAO 56 years). For country-specific distribution of variants, see https://www.mdsgene.org/d/41/g/4?action=plot_map&fc=0&_mu=1&_country=1. A positive family history was reported for almost a third (31.1%) of the PD patients with *LRRK2* variants, it was negative for 21.5% and unknown or not reported for 47.4%.

Information on age at onset (AAO) was available for 1,005 included patients (30.5%). The median age at onset (AAO) was 56 (range 20-95, interquartile range: 47-64) years (Figure 5A, B). While the majority of the patients (67.4%; n=677) had a late onset (≥50 years), approximately one-third (32.5%; n=327) had early-onset PD (EOPD), defined as an AAO after 21 and before 50 years of age.^46^ Notably, 9.3% (n=93) of included patients with information even had an AAO <40 years. Although one patient was reported with an AAO of 10 years^47^, it was actually 25 years (personal communication of Dr Lanza). The median disease duration among the included patients was 10 (range 0-42) years. When analyzing only carriers of VUS, the median AAO was 52.5 (range 20-79) years, suggesting that at least a portion of the VUS might also act as a driver of the disease since this AAO is younger than that reported for PD in general.^48^ For the group of patients with (likely) pathogenic variants, the median AAO was 56 years (the same as for all included patients, probably due to the high proportion of VUS carriers); the AAO range was 24-95 years. The median AAO of p.G2019S carriers was 57 (range 24-95) years (Figure 5B, Suppl. Table 5).

**Figure 5:**
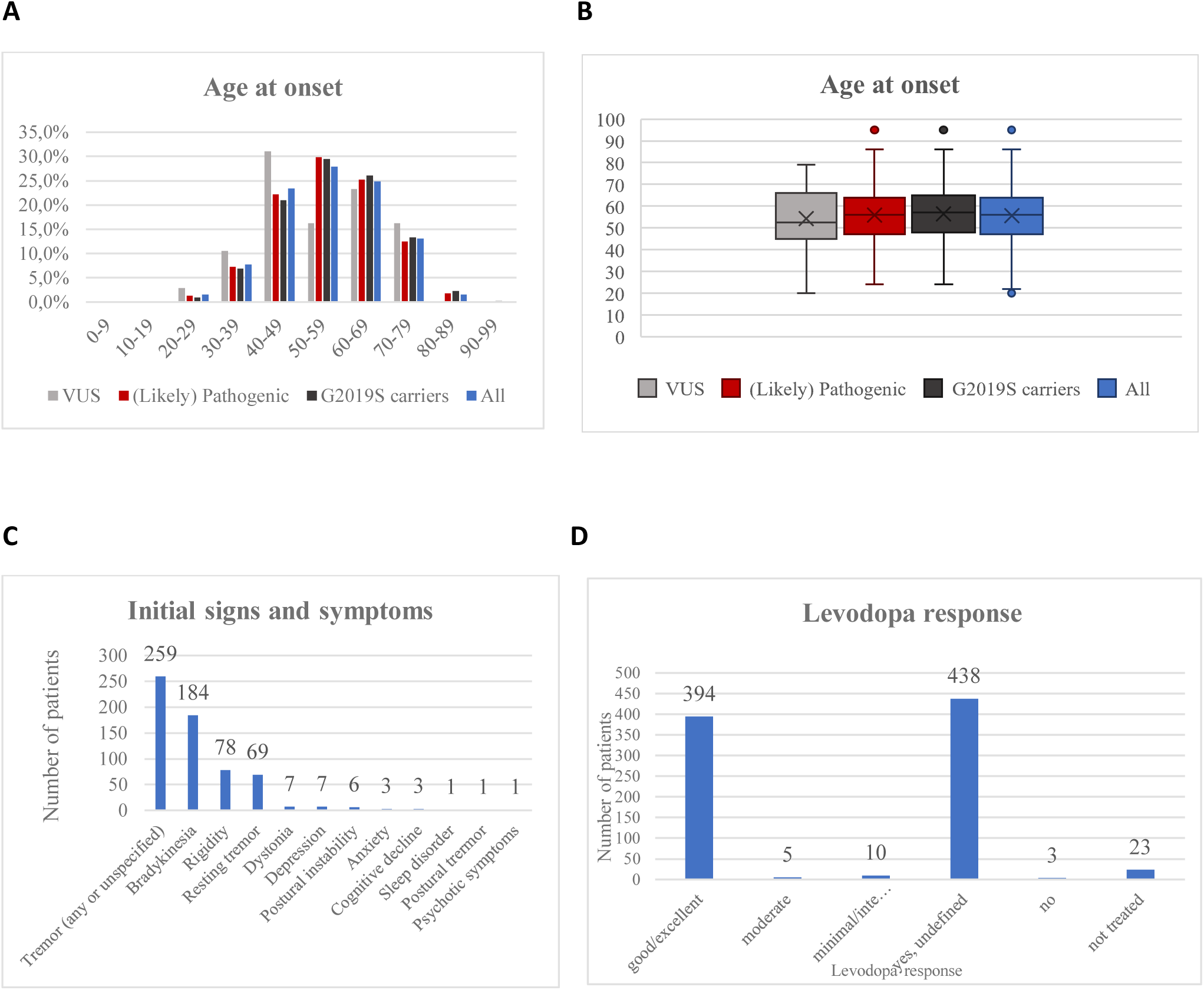
Other clinical features in the included patients. **A)** The age at onset (AAO) distribution is shown in 10-year bands on the x-axis. The number of patients is displayed on the y-axis. The graph shows the distribution for all included PD patients (blue), for the patients with VUS only (grey), and for patients with (likely) pathogenic variants (red). **B)** Box plots for the AAO in the three groups, depicting medians and interquartile ranges. Outlier points are also displayed and are defined as data points that lie outside 1.5 times the interquartile range (IQR) from the lower or upper quartile boundary. **C)** Initial signs and symptoms in the included patients. **D)** Levodopa response quantifications in the included PD patients. The x-axis shows the six divisions of the levodopa response quantifications and the y-axis the number of patients.

The data extraction covered 14 motor and seven non-motor symptoms (Suppl. Table 2). Motor symptoms were present in all included patients (per inclusion criterion), while at least one non-motor symptom was present in 9.4%, absent in 0.8%, and information was missing for 89.7% of the included patients. The most frequently reported motor symptom was bradykinesia, which was indicated as present in 74.4% of the patients and absent in 0.3% (according to the authors, who nevertheless established a diagnosis of PD). This was followed by rigidity (present in 21.0%, absent in 0.3%) and resting tremor (present in 12.4%, absent in 2.3%). Postural instability was the least reported of the four cardinal clinical features and was reported to be present in only 7.8% of the patients (Figure 4A). Among the non-cardinal motor symptoms, dyskinesia (present in 6.5%; absent in 3.1%; median disease duration in patients with dyskinesias: 14 years), motor fluctuations (present in 4.4%; absent in 2.1%; median disease duration in patients with motor fluctuations: 14 years), and dystonia (present in 2.1%; absent in 3.4%) were still relatively frequently reported, but the others were only reported to be present in less than 1% of the included patients with missing data in up to 99.9%, so that no meaningful conclusions can be drawn (Figure 4B). Among the non-motor symptoms, cognitive decline was the most frequently reported, present in 3.6% of the patients (median disease duration 11 years), followed by depression (present in 3.5%, median disease duration 11 years) (Suppl. Table 6). There were no clear differences regarding the frequency of motor and non-motor features in relation to the interpretation of the variant, i.e. (likely) pathogenic (85% of this group were carriers of p.G2019S) vs. VUS (Suppl. Table 7-8, Suppl. Figure 2).

Initial signs and symptoms were reported for 14.9% of the patients only. The most common initial symptom was tremor, which was present in 7.9% of all included patients and in 52.7% of the patients for whom information was available (Figure 5C). This was followed by bradykinesia (in 5.6% of all included patients; in 37.5% of the patients for whom information was available) and rigidity (in 2.4% of all included patients; in 15.9% of the patients for whom information was available). No clear differences were observed between carriers of variants considered (likely) pathogenic and VUS, except that resting tremor in carriers of VUS could be more frequent than in carriers of (likely) pathogenic variants (Suppl. Table 9, Suppl. Figure 3). Further subgroup analyses, for instance, for individual variants, were not meaningful due to the small number of carriers and high degree of missing data.

When comparing features of PARK-LRRK2 to other dominantly inherited forms of PD, i.e., PARK-SNCA or PARK-VPS35, it was noticeable that for all analyzed clinical features, *SNCA* and *VPS35* variant carriers had a higher proportion of symptoms reported as present than *LRRK2* variant carriers (www.mdsgene.org). In terms of initial presentation, the only difference found between PARK-LRRK2, PARK-SNCA, and PARK-VPS35 was that the most common sign and symptom in both PARK-SNCA (31.2%) and PARK-VPS35 patients (15.9%) was bradykinesia, while in *LRRK2* variant carriers, it was tremor (in 52.7% of the patients for whom information was available). Apart from tremor as an initial symptom in PARK-*LRRK2*, the identification of other clinical clues to the possible presence of PARK-LRRK2 was hampered by the high proportion of missing information in the published literature and needs to be an area of ongoing and future work.

For 26.5% of the patients (n=873, missing information in 73.5%), information was available on whether levodopa treatment was administered (yes for n=850). No information on the treatment response was provided for about half of them (51.5%). For those patients with information (n=412), almost all (95.6%) had a good/excellent response to levodopa (Figure 5D), with no apparent difference between carriers of (likely) pathogenic variants and VUS (Suppl. Table 10, Suppl. Figure 4). Information on other therapies, such as pump therapy or surgical treatment, including deep brain stimulation (DBS), was rarely available^49^ and not systematically analyzed. Reports by other investigators^50, 51^, including pooled analyses^52, 53^, indicate a beneficial effect of DBS in PARK-LRRK2.

## Discussion

This systematic literature review provides a comprehensive and up-to-date review of demographic, clinical, and genetic findings from the published literature on PARK-LRRK2. It is based on curated data from 3,296 patients with 205 different potentially disease-causing variants in *LRRK2*. This represents, by far, the largest database on PARK-LRRK2 to date. A comprehensive overview of *LRRK2* variants and interpretation is of high relevance to clinicians in establishing a possible diagnosis of PARK-LRRK2. This becomes increasingly actionable in light of the identification of several LRRK2 kinase inhibitors (MLi-2 (Merck), DNL-201, DNL-151/BIIB122 (Genentech/Denali), and PF-360 (Pfizer)), including first clinical trials for DNL-201 and DNL-151/BIIB122.^54^ The frequent discrepancies between variant interpretation using two automated online tools (i.e., Franklin and Varsome) highlight the value of an expert panel-guided variant interpretation for clinicians and genetic counselors.

Such a comprehensive review can help to identify clinical features to differentiate PARK-LRRK2 from idiopathic PD and other monogenic forms, at least on a group level, since the heterogeneity of the disease, for instance, the AAO ranging widely from 20-95 years, hampers the applicability at the individual level. Notably, the median AAO for PARK-LRRK2 in this review was 56 years, which is lower than the median AAO of 60-75 years reported for PD patients in general^48^, consistent with the notion that monogenic causes for PD typically have earlier AAO.^55^ Notably, around one-third of patients had EOPD with onset below age 50 years, thus adding nuance to a prevailing view that *LRRK2* variants cause late-onset PD^56, 57^. Compared to other genes linked to autosomal dominantly inherited PD, PARK-LRRK2 has a later median AAO vs. PARK-SNCA (46 years) but a similar AAO to PARK-VPS35 (52 years) patients (www.mdsgene.org). Genetic forms linked to autosomal recessive PD (*PARK7, PINK1, PRKN*) show even earlier median AAO (27, 31, 32 years; www.mdsgene.org).

In this systematic MDSGene review, we included, for the first time, patients with group-level data, i.e., publications in which groups of patients were reported with demographic and clinical data provided in aggregate (as means or percentages for the group but not as individual data). These patients account for 59.2% of the included patients. The disadvantage of the group level data is that they do not contain detailed individual clinical data and thus largely contribute to the high amount of missing data.

Therefore, the major challenge for this systematic literature review was the proportion of missing clinical data. Missingness of demographic and clinical features, such as AAO (information missing in ∼70% of patients) and ethnicity (missing in 50%), is an alarming observation. Likewise, the four cardinal motor features were unreported for 25.3% - 89.0% of patients, and information for other motor and non-motor signs and symptoms was even more frequently missing (ranging from 78.6% to 99.9%). This could be linked to the assumption that the authors of the papers with missing data implied the presence of certain symptoms (e.g., the cardinal features) or the absence of symptoms that are only rarely observed, or did not pay attention to a feature (absence of an examination).^3^ Despite these caveats, the reported prevalences of motor response complications and cognitive impairment in PARK-LRRK2 appeared lower in comparison to idiopathic PD. Dyskinesias and motor fluctuations were present in 6.5% and 4.4% of PARK-LRRK2 patients, respectively (with a median disease duration of 14 years), which were quite low compared with the widely quoted estimate of ∼10% of levodopa-treated PD patients per year developing motor response complications^45, 58, 59^. Similarly, although cognitive decline was the most frequently reported non-motor symptom in this systematic review, it was reportedly present in only 3.6% of patients (median disease duration 11 years), which was considerably lower than most published studies of idiopathic PD (for example, a recent review estimated that ∼20% of PD patients have cognitive impairment at the time of diagnosis, although their overall older age [mean 71.3±7.5 years, vs. 65.9±12.2 years in our study) is likely to have contributed to the higher rate of cognitive impairment.^60^ These observations are in line with studies comparing PARK-LRRK2 vs. idiopathic PD that, importantly, have adjusted for multiple potential confounders, including age, disease duration, and levodopa-equivalent dosages.^56, 61, 62^

In this work, a total of 292 *LRRK2* variants were documented in patients with different diseases, and 205 were included and classified as pathogenic, likely pathogenic, or VUS according to the ACMG criteria in patients with PD. Looking at the pathogenicity classifications, another challenge became obvious. As can be seen in Figure 2, the distribution of pathogenicity classifications using a commonly applied tool, i.e., Varsome was quite different from the manually curated one. Varsome often favored the classification of VUS as “likely benign,” which would mean that the patient/variant is not included in MDSGene and not considered for clinical trials. The observed discrepancies underline the need for careful checking of outputs from automated scoring systems by experts in the field^63^. In the manual pathogenicity scoring, all rare variants with a high CADD score were classified as VUS because it cannot be entirely ruled out that they do not affect the development of the disease. However, manual pathogenicity scoring according to ACMG is also challenging, as the criteria for interpretation are subjective and lead to classification inconsistencies.^64^ Importantly, the pathogenicity score is only an estimate and based on current knowledge, which may change over time as new insights become available. For example, variants currently classified as VUS may at some point be considered (likely) pathogenic if they are found to segregate in additional families and/or are demonstrated to have relevant functional/biological effects (e.g., elevated LRRK2 kinase activity). Currently, functional evidence is factored into pathogenicity scoring for a few variants only, since for most variants, little research has been done. One notable example is the novel p.F1700L variant in *LRRK2* with functional support of pathogenicity.^17^ Therefore, research studies that examine the impact of different *LRRK2* variants on kinase activity on a large scale^16^, are highly important to the field and are currently being generated.^65^

Correct variant interpretation has great relevance for the application of possible therapies, such as kinase inhibitors for PARK-LRRK2, because only patients who carry a kinase-activating *LRRK2* variant (and perhaps also sporadic PD patients who have biomarker evidence of elevated LRRK2 kinase activity) will likely respond to kinase inhibitors. Ultimately, understanding the functional impact of specific variants, which, given LRRK2’s wide-ranging physiological roles, may extend beyond the effects on kinase activity, is of great importance for the development of novel therapies.^66, 67^

Notably, an increasing number of publications suggest that *LRRK2* has a broader role beyond its link to classical PD. This includes its potential, albeit probably rare^36^, involvement in the pathogenesis of atypical parkinsonian disorders, as underlined by only <20 reported patients with potentially pathogenic variants in *LRRK2*, including MSA^29^ ^30, 68, 69^, PSP^21, 31–33, 68, 70^, CBD^31, 34^, and DLB^21^ patients. Of note, most of these patients carry only VUS (11/18). The possible role of *LRRK2* variants in atypical parkinsonian disorders accords with early observations of a broad pathological spectrum in the brains of patients with *LRRK2* variants, involving aggregation of alpha-synuclein and tau proteins.^71–73^ Importantly, long-term follow-up of patients initially diagnosed with PD may reveal additional patients with atypical parkinsonian disorders based on the subsequent clinical course or autopsy results, as illustrated by a patient reported as PD^74^, but later found to have MSA-P^75^. A further link between *LRRK2* and atypical parkinsonian disorders comes from a recently published genome-wide association study (GWAS) involving 1,001 White European-ancestry patients, which suggested a role of common *LRRK2* variation in the survival of PSP patients.^76^ Other phenotypes that have been associated with *LRRK2* variants in the literature include essential tremor, where a common variant might act as a risk factor in Asia^77^. Interestingly, *LRRK2* also has a role outside neurological disorders and has repeatedly been linked to inflammatory diseases.^78^ Notably, *LRRK2* shows its highest expression in blood and lung, the former also explaining why the functional testing of LRRK2’s kinase activity^79^ is possible in certain blood cells. A significantly higher incidence of inflammatory diseases like multiple sclerosis and rheumatoid arthritis has been reported^41^ and *LRRK2* postulated to be a link between gut inflammation and PD^67, 80^. Several studies have also shown that LRRK2 is functionally involved in infections with *Mycobacterium tuberculosis*^81, 82^ and *Mycobacterium leprae*^83, 84^. The observation of several somatic loss-of-function variants in *LRRK2* in breast cancer cells^42^ and germline variants in malignant mesothelioma^85^ are also interesting. Notably, patients with non-PD phenotypes often carried VUS or even benign variants, making their disease-causing role, in the sense of a monogenic disease, less clear and warrants further investigation.

Our study had some limitations, including, the unavailability of relevant clinical phenotype information in many cases. For instance, the presence or absence of sleep disorder was reported in only 3.9% of cases, making it challenging to conclude whether rapid eye movement sleep behavior disorder (RBD) - increasingly recognized to be a predictor of poorer prognosis in PD^86^ - is less common in PARK-LRRK2 compared to idiopathic PD, as suggested by some authors^56, 87^. Data on impulsive-compulsive behaviors, which are also of clinical relevance, similarly have rarely been reported and, thus, not been collected as part of MDSGene efforts. They need to be described more systematically and will be included in the future. Importantly, they will need to be contextualized with regards to patients’ treatment regimes, since dopamine agonist therapy is a primary risk factor for the occurrence of these behaviors. Further, the present MDSGene review can only deal with information at the time of reporting. The lack of confirmation of PD diagnosis by autopsy results and long-term clinical course that are highly relevant^88, 89^ is a limitation of this literature-based study. Therefore, dedicated studies are needed to evaluate the role of pathogenic *LRRK2* variants in the development of atypical parkinsonian features. Finally, modifiers of disease penetrance and AAO are areas of major interest but were beyond the scope of this review; for these issues, readers are referred to published works that have examined the effects of other genetic (including polygenic)^90, 91^, ethno-geographic^92^, and environmental factors^93^, and the interactions thereof^94^. We also did not assess the possible protective effect of LRRK2 p.G2019S among *GBA1* variant carriers^95–97^, which is an intriguing observation that opens up new questions and avenues for research.

In conclusion, we performed a systematic literature review, analyzed the data, and present insights from the largest database on PARK-LRRK2 to date. This review can be used to identify pathogenic variants and elucidate their demographic and phenotypic spectrum. Different filter options are available on the MDSGene website (https://www.mdsgene.org/), which also provides published information on in-vivo and in-vitro measurements of LRRK2 kinase activity. This database provides an important resource, especially in light of the emerging molecular-based therapies. However, the missing data, especially detailed clinical information, in the publications and the current limitations in variant interpretation, i.e., the high number of VUS, are important challenges that must be addressed to enable optimal selection and stratification of patients in ongoing and future clinical trials. Thus, this review contributes to improving precision medicine in PARK-LRRK2 patients.

## Data Availability

All data produced in the present study are available upon reasonable request to the authors.

https://www.mdsgene.org/

## Acknowledgments

This work was supported by the International Parkinson and Movement Disorder Society, the Parkinsońs Foundation, and the University of Luebeck. Financial disclosure statement: S.Y.L. has received grants from and served as a consultant for the Michael J. Fox Foundation. C.K. serves as a medical advisor to Centogene and Takeda and previously to Retromer Therapeutics and has received Speakers’ honoraria from Desitin and Bial. C.M. has received honoraria from the Parkinson’s Foundation.

## Notes

### Competing Interest Statement

The authors do not have any conflict of interest. S.Y.L. has received grants from and served as a consultant for the Michael J. Fox Foundation. C.K. serves as a medical advisor to Centogene and Takeda and previously to Retromer Therapeutics and has received Speakers' honoraria from Desitin and Bial. C.M. has received honoraria from the Parkinson's Foundation.

### Funding Statement

This work was supported by the International Parkinson and Movement Disorder Society, the Parkinson's Foundation, and the University of Luebeck.

### Author Declarations

The source data of our review article originated from published literature and were obtained through a systematic literature review using PubMed (https://pubmed.ncbi.nlm.nih.gov/).

