## Supplementary Tables and Figures for "MDSGene update and expansion: Clinical and genetic spectrum of *LRRK2* variants in Parkinson’s disease"

### Supporting Information

***Supplementary Table 1:*** Search term for the literature search for *LRRK2* in PubMed

| | |
|---|---|
| *LRRK2* | (ataxia OR ataxic OR cerebellar OR channelopathy OR dystonia OR dystonic OR parkinson* OR paroxysmal movement OR tremor OR myoclon* OR chorea OR choreo* OR choreatic OR spastic paraplegia OR spastic paraparesis OR HSP OR Strümpell* OR hyperkinetic OR “movement disorder” OR dyskinesia OR dyskinetic) AND (LRRK2 OR PARK8 OR DARDARIN OR RIPK7 OR ROCO2 OR AURA17 OR DKFZp434H2111 OR FLJ45829 OR RIPK7 OR “Leucin-rich repeat kinase 2” OR 12q12) AND ("english"[Language]) |

***Supplementary Table 2:*** List of variables. Demographic, clinical, and genetic information extracted using the MDSGene protocol.

| **Patient characterization** |
|---|
| individual patient ID |
| family ID |
| **Demographic information** |
| ethnicity |
| country of origin |
| sex |
| **Genetic information** |
| genetic status of all tested family members with number of homozygous, heterozygous and wildtype ones in affected and unaffected state |
| human genome version number |
| genome build or accession ID |
| gene to which the variant refers |
| physical location of the variant (on the plus strand) |
| observed mutated bases (on the plus strand) |
| reference bases (on the plus strand) |
| genomic description of the variant according to the nomenclature of the human genome variation society (HGVS) |
| coding description of the variant according to the nomenclature of the HGVS |
| protein description of the variant according to the nomenclature of the HGVS |
| genotype of the respective variant |
| type of variant |
| highest minor allele frequency (MAF) of the respective variant found in gnomAD |
| pathogenicity status classified as likely benign, benign, VUS (variant of unknown significance), likely pathogenic or pathogenic |
| CADD score as one parameter for pathogenicity scoring |
| Functional evidence |
| **Clinical information** |
| general |
| positive family history for the variant associated phenotype |
| age at examination in years |
| age at disease onset in years |
| disease duration in years |
| age at clinical diagnosis in years |
| age at death |
| presence of parkinsonian syndrome |
| motor instrument like UPDRS III or Hoehn&Yahr and, if available, their scoring |
| presence of non-motor signs and symptoms associated with a parkinsonian syndrome |
| non-motor symptoms instrument |
| presence of atypical signs and symptoms for a parkinsonian syndrome |
| reported initial signs and symptoms |
| motor signs and symptoms |
| bradykinesia |
| tremor (in general: HP:0001337) |
| rest tremor |
| action tremor |
| postural tremor |
| dystonic tremor |
| rigidity |
| postural instability |
| levodopa responsiveness |
| levodopa response quantification classified as minimal/intermittent, moderate, or good/excellent |
| dyskinesia |

| |
|---|
| dystonia |
| hyperreflexia |
| diurnal fluctuations as motor symptom fluctuations throughout the day |
| sleep benefit as restoration of mobility upon awakening from sleep |
| motor fluctuations in context of medication-intake |
| non-motor signs and symptoms |
| hyposmia |
| depression |
| depression scale |
| anxiety |
| anxiety scale |
| psychotic symptoms |
| psychosis scale |
| sleep disorder |
| cognitive decline |
| autonomic symptoms |

***Supplementary Table 3***: Documented *LRRK2* variants in all extracted variant carriers.

**A:** Variants classified as pathogenic, likely pathogenic, likely benign, or benign

| Pathogenic | Likely Pathogenic | Likely Benign | Benign |
|---|---|---|---|
| R1325Q | R767H | H115P | IVS2+1366C>G |
| N1437H | R1067Q | L119P | IVS5+33T>C |
| R1441C | N1437D | IVS4-38A>G | N551K |
| R1441G | N1437S | M262V | I723V |
| R1441H | F1700L | P755L | IVS19-10dupT |
| R1441S | S1761R | S865F | Q1111H |
| A1442P | S1954F[1] | C925Y | P1262A |
| V1447M | I2020L | L1281L | R1398H |
| Y1699C | I2020S | I1371V | T1410M |
| F1700L | T2031S | R1441R | K1423K |
| L1795F[2] | | Y1527Y | IVS30+12delT |
| G2019S | | Y2018Y | R1514Q |
| I2020T | | H2391Q | M1646T |
| | | | S1647T |
| | | | N2081D |
| | | | G2385G |

[1] This variant was only found in a patient with breast cancer and not included in MDSGene

[2] This variant is encoded by two different substitutions on the cDNA level (c.5385G>C and c.5385G>T)

**B:** Variants classified as VUS

| | | | | | |
|---|---|---|---|---|---|
| E10K | K544E | S1007T | S1445F | Y1649S | E2230K |
| K53R | M579V | H1019R | P1446L | P1661T | H2236R |
| M100T | I610T | Q1029H | V1450I | R1677S | N2251T |
| Q116R | K616R | N1089S | G1451S | I1691T | N2261K |
| S128G | S633F | S1096C | A1464G | R1693Q | G2294R |
| L153W | T646A | L1114L | K1468E | S1706* | N2308D |
| N173S | A654V | I1122V | P1480L | R1707K | T2310M |
| S192A | S661F | L1128M | A1481T | L1712F | N2313S |
| E193A | I675M | A1151T | R1483Q | R1725Q | I2323F |
| E193K | F676S | C1152F | A1490V | S1752C | I2336V |
| A211V | S722N | S1159R | T1491del | D1756G | S2350I |
| C228S | M724V | L1165P | E1492K | D1756Y | T2356I |
| H230R | D734N | S1181Y | R1501W | H1758P | G2385R# |
| N238I | K739R | I1192M | I1505Rfs*16 | R1771T | V2390A |
| Y250C | S764N | I1192V | S1508G | K1772del | V2390M |
| E255K | R767C | A1215T | G1520A | C1774Y | E2395K |
| K258N | R772* | H1216R | V1541M | T1806I | M2408I |
| A259E | T776M | S1228T | I1548V | G1819R | T2423S |
| L286V | I777M | T1271I | R1552* | Q1823K | I2434V |
| V291A | S784R | W1295R | A1589S | M1869T | L2439I |
| A306V | I786F | L1304F | V1613A | M1869V | A2461V |
| L308M | R792K | R1334Q | V1615M | E1874* | L2463P |
| A312V | R793M | I1339M | V1615V | D1887N | Q2490H |
| T320S | S827F | V1340M | E1616K | G1891A | T2494I |
| S231P | I844N | Q1353K | P1619L | L1914I | V2495I |
| E334K# | M848T | V1369A | K1620R | R1941H | T2524A |
| N335S | S858* | V1373M | H1621R | Q1961R | IVS4-68T>C |
| A370T | S885C | Q1379R | H1621Q | I1991V | IVS8+1G>A |
| L378F | E899D | L1388I | P1622L | Y2006H | IVS19+5delGTAA |
| M379I | L922* | V1389I | P1622P | A2010T | IVS30+1G>C |
| I388T | Q923H | R1398C | P1622T | I2012T | IVS31+3A>G |
| V414I | Q930R | R1398R | K1623E | L2063* | IVS33+6T>A |
| A419V | D941Y | Y1402C | I1626V | V2074I | IVS33+78T>C |
| L425P | D944V | H1405Q | S1627L | E2108K | IVS33+70T>C |
| L437S | R948Q | A1413T | S1627T | N2133S | IVS33+79T>C |
| S438P | S958L | R1320S | R1628C | C2139S | IVS36-29G>T |
| A459S | D972G | E1427G | R1628P# | V2150I | IVS40-28A>T |
| S461I | S973G | F1436L | R1639Kfs*13 | W2168L | IVS49+178A>G |
| N474S | S973N | A1440P | Y1645S | D2175H | Exon 49 deletion |
| R521G | R981K | S1445C | Q1648R | Y2189C | |
| K535Nfs*13 | | | | | |

#: established risk variants

***Supplementary Table 4***: Numbers and frequencies of pathogenic or likely pathogenic *LRRK2* variants in affected PD patients and unaffected individuals.

| Variant | Number of affected patients (frequency) | | Number of unaffected carriers |
|---|---|---|---|
| **Pathogenic variants** | | | |
| G2019S | 2426 | (73.60%) | 739 |
| R1441G | 193 | (5.86%) | 90 |
| R1441C | 96 | (2.91%) | 17 |
| I2020T | 43 | (1.30%) | 5 |
| R1441H | 39 | (1.18%) | 4 |
| Y1699C | 32 | (0.97%) | 0 |
| N1437H | 9 | (0.27%) | 1 |
| V1447M | 6 | (0.18%) | 0 |
| L1795F | 4 | (0.12%) | 0 |
| R1441S | 4 | (0.12%) | 0 |
| R1325Q | 3 | (0.09%) | 6 |
| F1700L | 2 | (0.06%) | 0 |
| A1442P | 1 | (0.03%) | 0 |
| **Likely pathogenic variants** | | | |
| R1067Q | 9 | (0.27%) | 0 |
| N1437D | 7 | (0.21%) | 0 |
| S1761R | 7 | (0.21%) | 0 |
| N1437S | 5 | (0.15%) | 0 |
| I2020L | 1 | (0.03%) | 0 |
| I2020S | 1 | (0.03%) | 0 |
| R767H | 1 | (0.03%) | 0 |
| T2031S | 1 | (0.03%) | 0 |
| S1954F* | 0 | (0.00%) | 0 |

* found in 1 patient with breast cancer

***Supplementary Table 5***: Age at onset (AAO) of PD in all included patients, in the included patients with VUS and in the included patients with (likely) pathogenic variants. First number indicates the number of patients, number in parentheses is the respective frequency.

| **Age at onset** | **Patients with VUS** | | **Patients with (likely) pathogenic variants** | | **All patients** | |
|---|---|---|---|---|---|---|
| 0-9 | 0 | (0.0%) | 0 | (0.0%) | 0 | (0.0%) |
| 10-19 | 0 | (0.0%) | 0 | (0.0%) | 0 | (0.0%) |
| 20-29 | 4 | (2.8%) | 11 | (1.3%) | 15 | (1.5%) |
| 30-39 | 15 | (10.6%) | 63 | (7.3%) | 78 | (7.8%) |
| 40-49 | 44 | (31.0%) | 191 | (22.1%) | 235 | (23.4%) |
| 50-59 | 23 | (16.2%) | 257 | (29.8%) | 280 | (27.9%) |
| 60-69 | 33 | (23.2%) | 217 | (25.1%) | 250 | (24.9%) |
| 70-79 | 23 | (16.2%) | 108 | (12.5%) | 131 | (13.0%) |
| 80-89 | 0 | (0.0%) | 15 | (1.7%) | 15 | (1.5%) |
| 90-99 | 0 | (0.0%) | 1 | (0.1%) | 1 | (0.1%) |
| known | 142 | (100.0%) | 863 | (100.0%) | 1005 | (100.0%) |
| unknown | 260 | | 2031 | | 2291 | |
| total | 402 | | 2894 | | 3296 | |

***Supplementary Table 6***: Information on reported signs and symptoms in all included patients. First number indicates the number of patients, number in parentheses is the respective frequency.

| **Symptoms** | **Present** | | **Absent** | | **Unknown** | |
|---|---|---|---|---|---|---|
| Motor symptoms | 3296 | (100%) | 0 | (0.0%) | 0 | (0.0%) |
| Parkinsonism | 3296 | (100%) | 0 | (0.0%) | 0 | (0.0%) |
| Bradykinesia | 2452 | (74.4%) | 10 | (0.3%) | 834 | (25.3%) |
| Rigidity | 692 | (21.0%) | 10 | (0.3%) | 2594 | (78.7%) |
| Tremor (any or unspecified) | 636 | (19.3%) | 70 | (2.1%) | 2590 | (78.6%) |
| Resting tremor | 409 | (12.4%) | 75 | (2.3%) | 2812 | (85.3%) |
| Action tremor | 20 | (0.6%) | 7 | (0.2%) | 3269 | (99.2%) |
| Postural tremor | 24 | (0.7%) | 10 | (0.3%) | 3262 | (99.0%) |
| Dystonic tremor | 0 | (0.0%) | 3 | (0.1%) | 3293 | (99.9%) |
| Postural instability | 258 | (7.8%) | 104 | (3.2%) | 2934 | (89.0%) |
| Dyskinesia | 213 | (6.5%) | 103 | (3.1%) | 2980 | (90.4%) |
| Dystonia | 70 | (2.1%) | 111 | (3.4%) | 3115 | (94.5%) |
| Hyperreflexia | 12 | (0.4%) | 49 | (1.5%) | 3235 | (98.1%) |
| Diurnal fluctuations | 2 | (0.1%) | 0 | (0.0%) | 3294 | (99.9%) |
| Sleep benefit | 17 | (0.5%) | 59 | (1.8%) | 3220 | (97.7%) |
| Motor flucttutations | 146 | (4.4%) | 68 | (2.1%) | 3082 | (93.5%) |
| Non-motor symptoms | 311 | (9.4%) | 28 | (0.8%) | 2957 | (89.7%) |
| Hyposmia | 46 | (1.4%) | 11 | (0.3%) | 3239 | (98.3%) |
| Depression | 114 | (3.5%) | 160 | (4.9%) | 3022 | (91.7%) |
| Anxiety | 49 | (1.5%) | 110 | (3.3%) | 3137 | (95.2%) |
| Psychotic symptoms | 67 | (2.0%) | 141 | (4.3%) | 3088 | (93.7%) |
| Sleep disorder | 72 | (2.2%) | 55 | (1.7%) | 3169 | (96.1%) |
| Cognitive decline | 120 | (3.6%) | 331 | (10.0%) | 2845 | (86.3%) |
| Autonomic symptoms | 63 | (1.9%) | 79 | (2.4%) | 3154 | (95.7%) |
| Atypical parkinsonism symptoms | 10 | (0.3%) | 311 | (9.4%) | 2975 | (90.3%) |

***Supplementary Table 7*:** Information on reported signs and symptoms in the patients with (likely) pathogenic variants (n=2,894). First number indicates the number of patients, number in parentheses is the respective frequency.

| Symptoms | Present | | Absent | | Unknown | |
|---|---|---|---|---|---|---|
| Motor symptoms | 2894 | (100.0%) | 0 | (0.0%) | 0 | (0.0%) |
| Parkinsonism | 2894 | (100.0%) | 0 | (0.0%) | 0 | (0.0%) |
| Bradykinesia | 2176 | (75.2%) | 8 | (0.3%) | 710 | (24.5%) |
| Rigidity | 621 | (21.5%) | 7 | (0.2%) | 2266 | (78.3%) |
| Tremor (any or unspecified) | 556 | (19.2%) | 63 | (2.2%) | 2275 | (78.6%) |
| Resting tremor | 352 | (12.2%) | 60 | (2.1%) | 2482 | (85.8%) |
| Action tremor | 17 | (0.6%) | 7 | (0.2%) | 2870 | (99.2%) |
| Postural tremor | 18 | (0.6%) | 9 | (0.3%) | 2867 | (99.1%) |
| Dystonic tremor | 0 | (0.0%) | 3 | (0.1%) | 2891 | (99.9%) |
| Postural instability | 230 | (7.9%) | 83 | (2.9%) | 2581 | (89.2%) |
| Dyskinesia | 201 | (6.9%) | 88 | (3.0%) | 2605 | (90.0%) |
| Dystonia | 65 | (2.2%) | 96 | (3.3%) | 2733 | (94.4%) |
| Hyperreflexia | 12 | (0.4%) | 46 | (1.6%) | 2836 | (98.0%) |
| Diurnal fluctuations | 2 | (0.1%) | 0 | (0.0%) | 2892 | (99.9%) |
| Sleep benefit | 15 | (0.5%) | 59 | (2.0%) | 2820 | (97.4%) |
| Motor flucttutations | 135 | (4.7%) | 65 | (2.2%) | 2694 | (93.1%) |
| Non-motor symptoms | 265 | (9.2%) | 28 | (1.0%) | 2601 | (89.9%) |
| Hyposmia | 37 | (1.3%) | 7 | (0.2%) | 2850 | (98.5%) |
| Depression | 104 | (3.6%) | 150 | (5.2%) | 2640 | (91.2%) |
| Anxiety | 47 | (1.6%) | 110 | (3.8%) | 2737 | (94.6%) |
| Psychotic symptoms | 63 | (2.2%) | 133 | (4.6%) | 2698 | (93.2%) |
| Sleep disorder | 60 | (2.1%) | 48 | (1.7%) | 2786 | (96.3%) |
| Cognitive decline | 105 | (3.6%) | 311 | (10.7%) | 2478 | (85.6%) |
| Autonomic symptoms | 51 | (1.8%) | 74 | (2.6%) | 2769 | (95.7%) |
| Atypical parkinsonism symptoms | 8 | (0.3%) | 305 | (10.5%) | 2581 | (89.2%) |

***Supplementary Table 8***: Information on reported signs and symptoms in the patients with VUS (n=402). First number indicates the number of patients, number in parentheses is the respective frequency.

| Symptoms | Present | | Absent | | Unknown | |
|---|---|---|---|---|---|---|
| Motor symptoms | 402 | (100.0%) | 0 | (0.0%) | 0 | (0.0%) |
| Parkinsonism | 402 | (100.0%) | 0 | (0.0%) | 0 | (0.0%) |
| Bradykinesia | 276 | (68.7%) | 2 | (0.5%) | 124 | (30.8%) |
| Rigidity | 71 | (17.7%) | 3 | (0.7%) | 328 | (81.6%) |
| Tremor (any or unspecified) | 80 | (19.9%) | 7 | (1.7%) | 315 | (78.4%) |
| Resting tremor | 57 | (14.2%) | 15 | (3.7%) | 330 | (82.1%) |
| Action tremor | 3 | (0.7%) | 0 | (0.0%) | 399 | (99.3%) |
| Postural tremor | 6 | (1.5%) | 1 | (0.2%) | 395 | (98.3%) |
| Dystonic tremor | 0 | (0.0%) | 0 | (0.0%) | 402 | (100.0%) |
| Postural instability | 28 | (7.0%) | 21 | (5.2%) | 353 | (87.8%) |
| Dyskinesia | 12 | (3.0%) | 15 | (3.7%) | 375 | (93.3%) |
| Dystonia | 5 | (1.2%) | 15 | (3.7%) | 382 | (95.0%) |
| Hyperreflexia | 0 | (0.0%) | 3 | (0.7%) | 399 | (99.3%) |
| Diurnal fluctuations | 0 | (0.0%) | 0 | (0.0%) | 402 | (100.0%) |
| Sleep benefit | 2 | (0.5%) | 0 | (0.0%) | 400 | (99.5%) |
| Motor fluctutations | 10 | (2.5%) | 3 | (0.7%) | 389 | (96.8%) |
| Non-motor symptoms | 43 | (10.7%) | 0 | (0.0%) | 359 | (89.3%) |
| Hyposmia | 9 | (2.2%) | 4 | (1.0%) | 389 | (96.8%) |
| Depression | 10 | (2.5%) | 10 | (2.5%) | 382 | (95.0%) |
| Anxiety | 2 | (0.5%) | 0 | (0.0%) | 400 | (99.5%) |
| Psychotic symptoms | 4 | (1.0%) | 8 | (2.0%) | 390 | (97.0%) |
| Sleep disorder | 12 | (3.0%) | 7 | (1.7%) | 383 | (95.3%) |
| Cognitive decline | 15 | (3.7%) | 19 | (4.7%) | 368 | (91.5%) |
| Autonomic symptoms | 12 | (3.0%) | 5 | (1.2%) | 385 | (95.8%) |
| Atypical parkinsonism symptoms | 2 | (0.5%) | 6 | (1.5%) | 394 | (98.0%) |

***Supplementary Table 9***: Reported initial signs and symptoms in the included patients with VUS, in patients with (likely) pathogenic variants, and in all included patients. First number indicates the number of patients, numbers in parentheses are the respective frequencies among all patients in the same group (middle panel) and patients in the same group with information (right panel).

| **Initial symptoms** | **Patients with VUS** | | | **Patients with (likely) pathogenic variants** | | | **All included patients** | | |
|---|---|---|---|---|---|---|---|---|---|
| Tremor (any or unspecified) | 19 | (4.8%) | (43.2%) | 240 | (8.3%) | (53.7%) | 259 | (7.9%) | (52.7%) |
| Bradykinesia | 16 | (4.0%) | (36.4%) | 168 | (5.8%) | (37.6%) | 184 | (5.6%) | (37.5%) |
| Rigidity | 5 | (1.2%) | (11.4%) | 73 | (2.5%) | (16.3%) | 78 | (2.4%) | (15.9%) |
| Resting tremor | 11 | (2.7%) | (25.0%) | 58 | (2.0%) | (13.0%) | 69 | (2.1%) | (14.1%) |
| Postural instability | 1 | (0.2%) | (2.3%) | 5 | (0.2%) | (1.1%) | 6 | (0.2%) | (1.2%) |
| Dystonia | 0 | (0.0%) | (0.0%) | 7 | (0.2%) | (1.6%) | 7 | (0.2%) | (1.4%) |
| Anxiety | 0 | (0.0%) | (0.0%) | 3 | (0.1%) | (0.7%) | 3 | (0.1%) | (0.6%) |
| Cognitive decline | 1 | (0.2%) | (2.3%) | 2 | (0.1%) | (0.4%) | 3 | (0.1%) | (0.6%) |
| Sleep disorder | 0 | (0.0%) | (0.0%) | 1 | (0.0%) | (0.2%) | 1 | (0.0%) | (0.2%) |
| Depression | 0 | (0.0%) | (0.0%) | 7 | (0.2%) | (1.6%) | 7 | (0.2%) | (1.4%) |
| Tremor postural | 0 | (0.0%) | (0.0%) | 1 | (0.0%) | (0.2%) | 1 | (0.0%) | (0.2%) |
| Psychotic symptoms | 0 | (0.0%) | (0.0%) | 1 | (0.0%) | (0.2%) | 1 | (0.0%) | (0.2%) |
| Known | 44 | (10.9%) | (100.0%) | 447 | (15.4%) | (100.0%) | 491 | (14.9%) | (100.0%) |
| Unknown | 358 | (89.1%) | | 2447 | (84.6%) | | 2805 | (85.1%) | |
| All | 402 | (100.0%) | | 2894 | (100.0%) | | 3296 | (100.0%) | |

***Supplementary Table 10*:** Levodopa response quantifications in the included PD patients with VUS, in the included PD patients with (likely) pathogenic *LRRK2* variants, and in all included PD patients.

| **Levodopa response** | **Patients with VUS** | | **Patients with (likely) pathogenic variants** | | **All included patients** | |
|---|---|---|---|---|---|---|
| good/excellent | 53 | (13.2%) | 341 | (11.8%) | 394 | (12.0%) |
| moderate | 0 | (0.0%) | 5 | (0.2%) | 5 | (0.2%) |
| minimal/intermittent | 2 | (0.5%) | 8 | (0.3%) | 10 | (0.3%) |
| yes, undefined | 34 | (8.5%) | 404 | (14.0%) | 438 | (13.3%) |
| no | 1 | (0.2%) | 2 | (0.1%) | 3 | (0.1%) |
| not treated | 5 | (1.2%) | 18 | (0.6%) | 23 | (0.7%) |
| unknown | 307 | (76.4%) | 2116 | (73.1%) | 2423 | (73.5%) |
| total | 402 | (100.0%) | 2894 | (100.0%) | 3296 | (100.0%) |

**A**

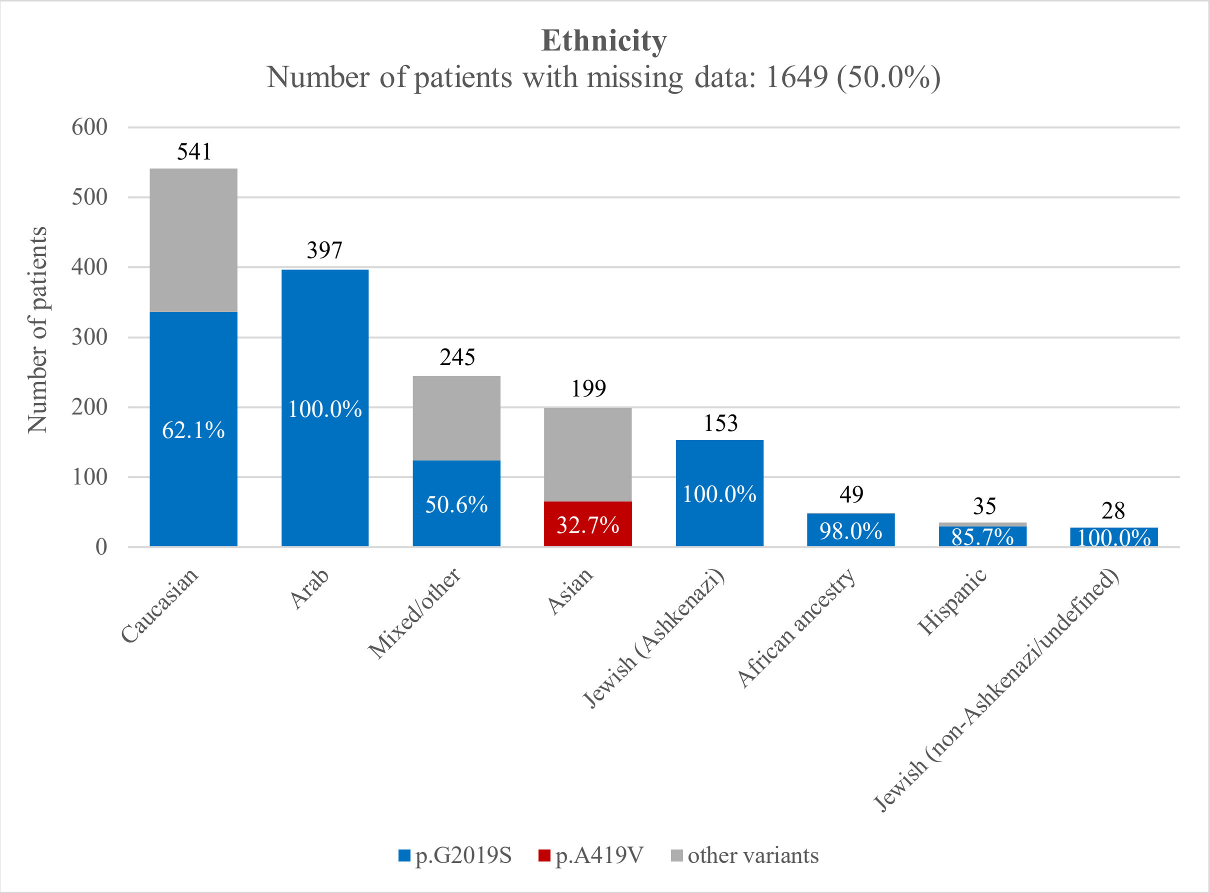

Ethnicity
Number of patients with missing data: 1649 (50.0%)
Number of patients
600
500
400
300
200
100
0
541
397
245
199
153
49
35
28
62.1%
100.0%
50.6%
32.7%
100.0%
98.0%
85.7%
100.0%
Caucasian
Arab
Mixed/other
Asian
Jewish (Ashkenazi)
African ancestry
Hispanic
Jewish (non-Ashkenazi/undefined)
p.G2019S
p.A419V
other variants

**B**

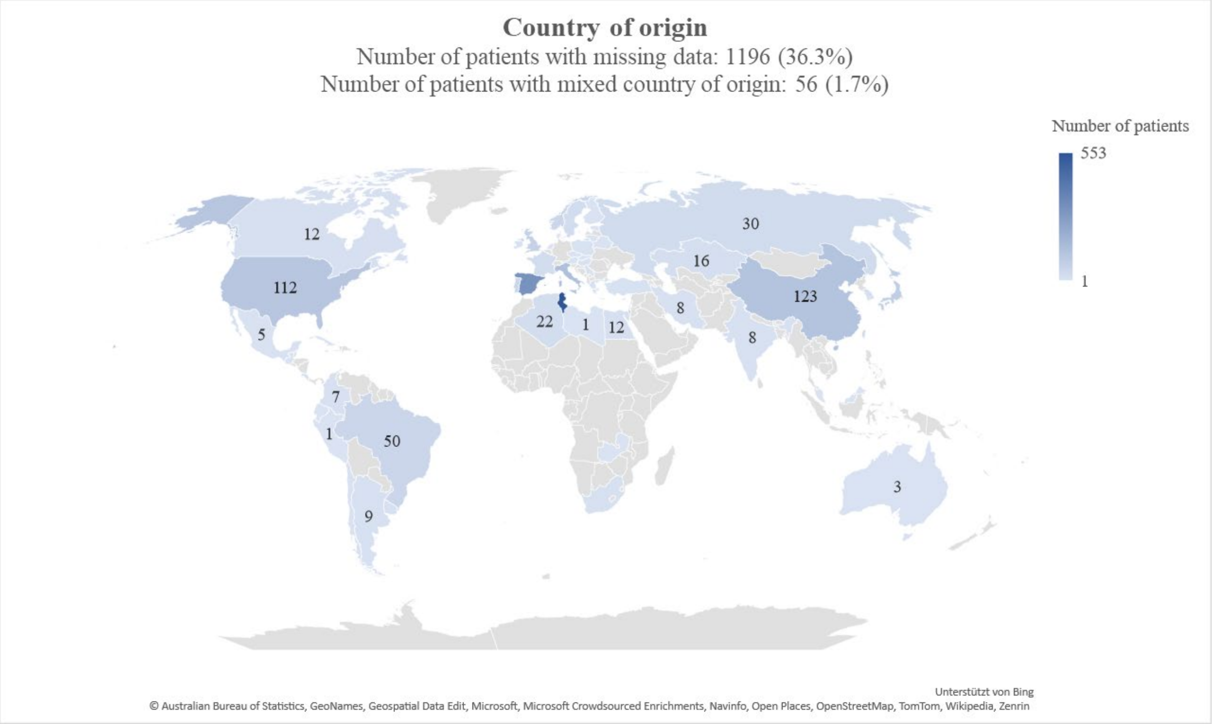

Country of origin
Number of patients with missing data: 1196 (36.3%)
Number of patients with mixed country of origin: 56 (1.7%)
Number of patients
553
1
12
112
5
30
16
123
8
8
22
1
12
7
1
50
9
3
Unterstützt von Bing
© Australian Bureau of Statistics, GeoNames, Geospatial Data Edit, Microsoft, Microsoft Crowdsourced Enrichments, Navinfo, Open Places, OpenStreetMap, TomTom, Wikipedia, Zenrin

***Supplementary Figure 1*****:** Origin of included PD patients. **A)** Ethnicities of the included patients with the most frequent variant per ethnicity highlighted. **B)** World map for included PD patients with *LRRK2* variants. The country of origin with the highest number of affected *LRRK2* carriers is Tunisia (n = 553, followed by Spain (n = 313) and Italy (n = 135).

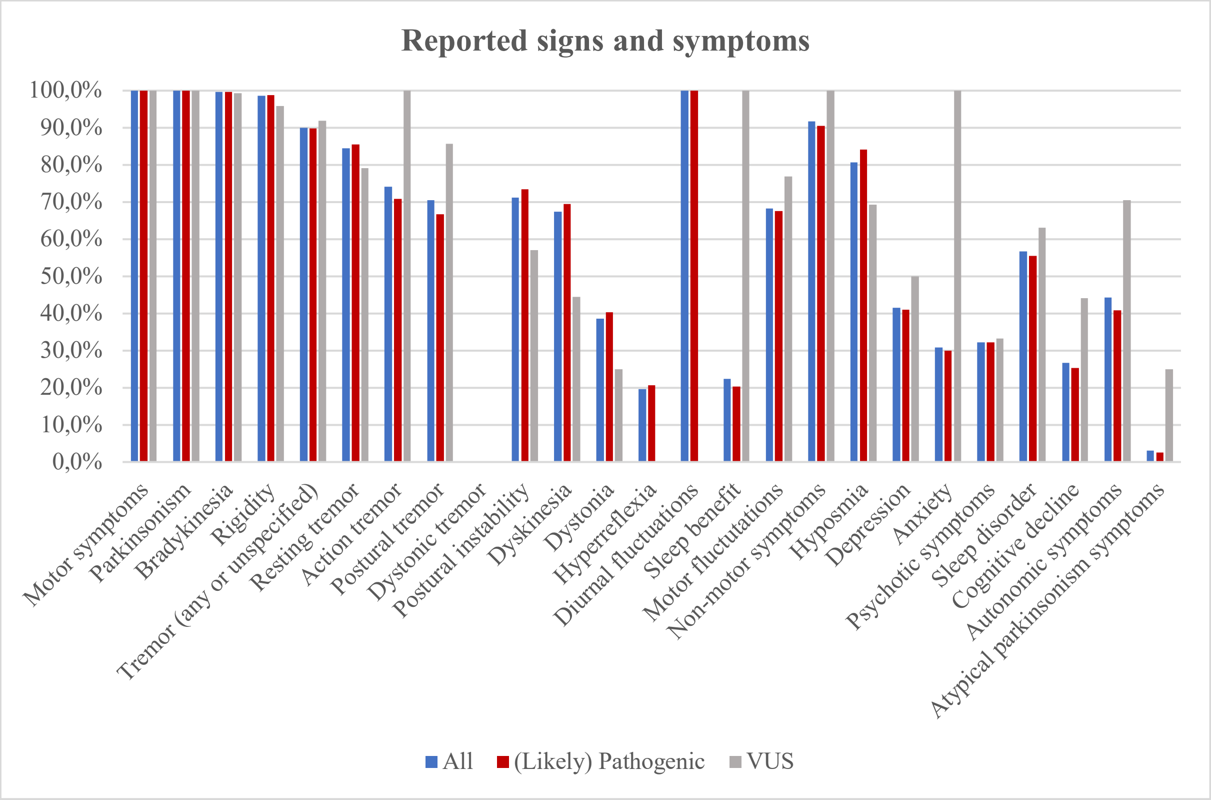

***Supplementary Figure 2*****:** Percentages of features reported to be present among all patients in the respective group. All reported signs and symptoms ("present") in the included patients with VUS (n=402), with (likely) pathogenic variants (n=2,894), and in all included PD patients (n=3,296). Of note, there is bias in this presentation since the diagram includes only information from patients with available data and there might be overreporting of unusual features combined with underreporting of the absence of respective features (missing data) leading to a too-high frequency, especially when the sample size (patients with information) is small.

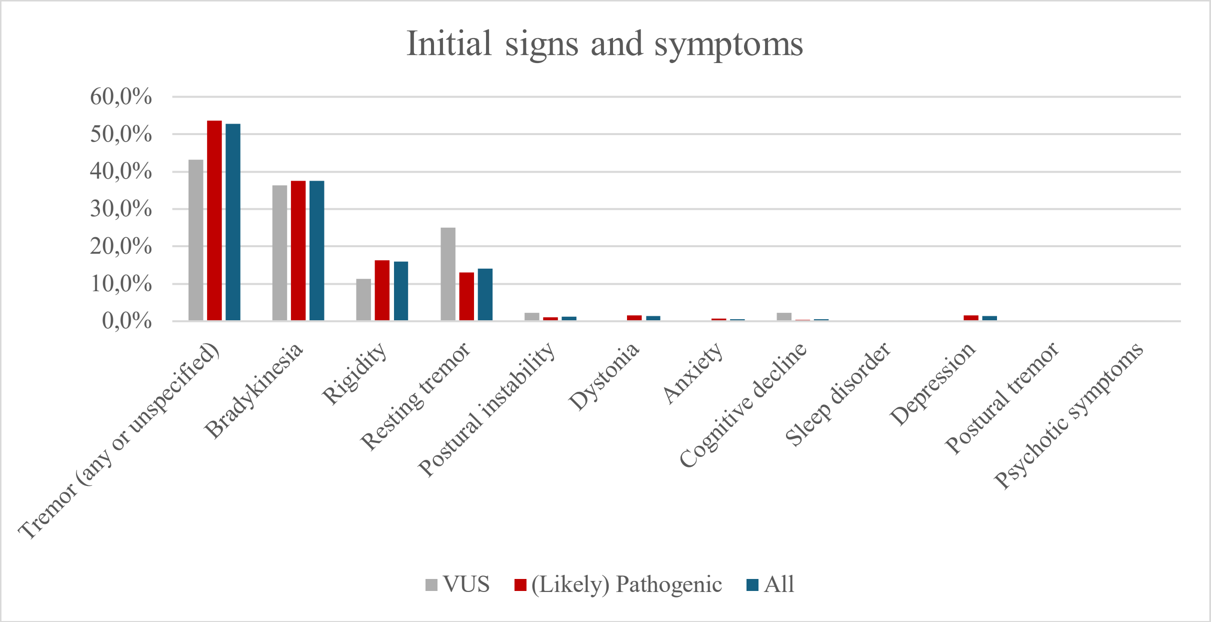

***Supplementary Figure 3*****:** Initial signs and symptoms ("present") in all included patients with information (blue, n=491), in the included patients with VUS (gray, n=44) and with (likely) pathogenic variants (red, n=447).

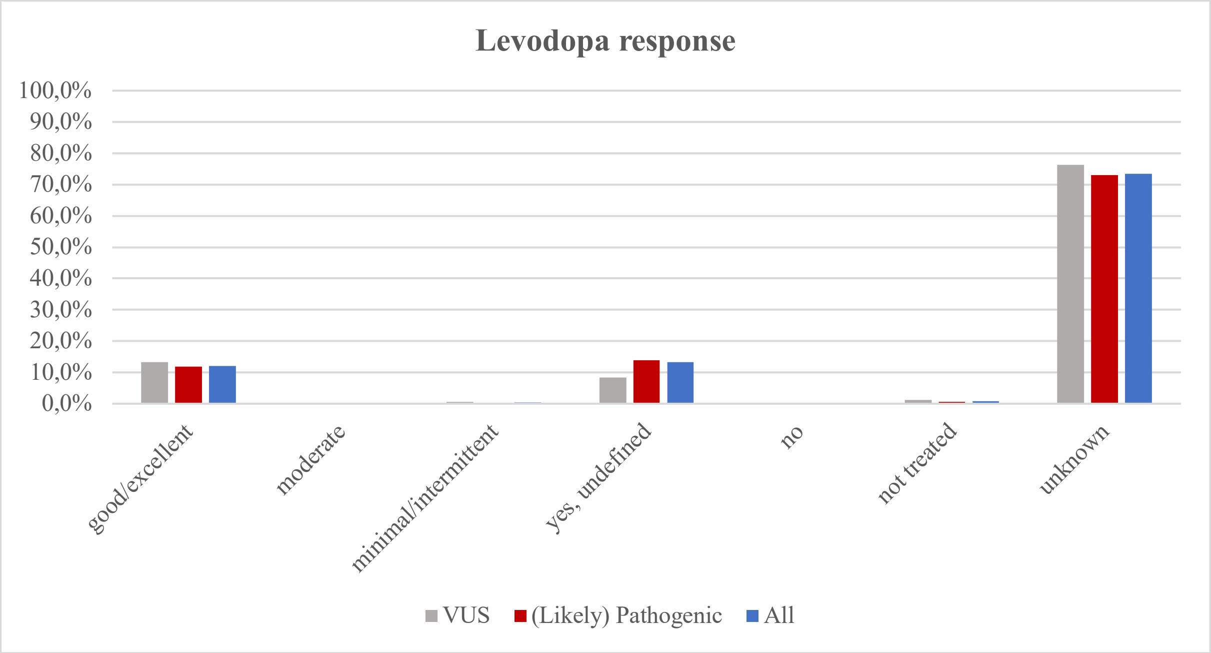

***Supplementary Figure 4*****:** Levodopa response quantifications in all included PD patients with information (blue, n=873), in the included PD patients with VUS (gray, n=95) and in the included PD patients with (likely) pathogenic *LRRK2* variants (red, n=778).
